# A Proof-of-Concept Clinical Trial Design for Evolutionary Guided Precision Medicine for Cancer

**DOI:** 10.1101/2025.05.23.25328210

**Authors:** Deepak Parashar, Wei He, Peter Mon, Martyn Plummer, Matthew D. McCoy, Chen-Hsiang Yeang, Robert A. Beckman

## Abstract

Current Precision Medicine (CPM) matches cancer therapies to consensus molecular characteristics at one or more timepoints. However, cancers contain extensive subclonal heterogeneity leading to dynamic evolution in response to therapy. Mathematical modeling has the potential to optimize timing and sequencing of therapies in a more effective and personalized manner than CPM. Clinical trial designs testing Evolutionary Guided Precision Medicine (EGPM) strategies for preventing or delaying relapse to improve outcomes, are needed. In a simulation, we evaluated Dynamic Precision Medicine (DPM), an EGPM, in a stratified randomized design based on whether the patient was predicted to benefit from DPM, using an evolutionary classifier. We present this new proof-of-concept clinical trial design and perform simulations which show high power, control of false positive rates, and robust performance in the face of anticipated challenges to clinical translation. The design is distinct from common biomarker-driven designs and can provide a robust evaluation of EGPM.

---

**Current precision medicine (CPM)** is a major direction in oncology and has substantially benefitted patients. It matches therapies to static, consensus molecular patterns in bulk samples. Patients are treated with a matching therapy for as long as they are deemed to be benefiting, i.e. until radiologic progression, clinical progression or relapse. The process is then repeated with a different therapy. In spite of the benefits of this approach, each repetition of this process typically results in diminishing returns. Long term responses and cures remain elusive.

These clinical results are in part the result of pre-existing genetic subclonal heterogeneity within cancers. A deep duplex sequencing [1] study of colorectal cancer at 20,000x depth and exceptional accuracy, and associated mathematical modelling indicate that, for any cancer large enough to be visible on computed tomography, every viable resistance mechanism to a single therapy will be pre-existing, albeit often in small undetectable subclones [2-4]. These studies not only indicate that intratumoral genetic diversity exceeds that previously acknowledged, but that evolution of cells with resistance to multiple non-cross resistant therapies in the same cell accelerates with increasing cancer load to an even greater extent than previously assumed. Moreover, as resistance is selected for, “hypermutator subclones” that evolve even more rapidly due to mutations affecting genome maintenance will also be selected, since they are more likely to have pre-existing resistance mutations and to acquire new mutations. These findings emphasize the inevitable challenge of clinical resistance and the increasing difficulty of providing clinical benefit later in a patient’s clinical course.

**Evolutionary guided precision medicine (EGPM)** approaches explicitly consider subclonal heterogeneity and evolutionary dynamics in an attempt to slow the evolution of resistance and prolong survival. A number of approaches are under development, some of which are described in references [5-9].

Our work is focused on **dynamic precision medicine (DPM)** [7], a particular approach to EGPM. DPM explicitly considers minority subclones and heritable genetic and epigenetic evolutionary dynamics, with the goal of treating current disease and preventing refractory disease relapse by blocking future acquisition of simultaneous resistance to multiple non-cross resistant agents. (Notably, many agents do exhibit cross resistance and DPM requires availability of at least two therapies with limited cross resistance). As a result, DPM will often recommend prioritizing elimination of rare hypermutator subclones vs. debulking the tumor. This involves administering brief pulses of later line therapies in earlier lines. Figure 1 illustrates the key underlying principles of DPM. DPM uses specialized decision criteria and a mathematical model to prioritize among two treatment priorities (immediate cytoreduction and resistance prevention) as a function of evolutionary dynamics, in contrast to CPM, which typically focuses on immediate cytoreduction.

**Figure 1.**
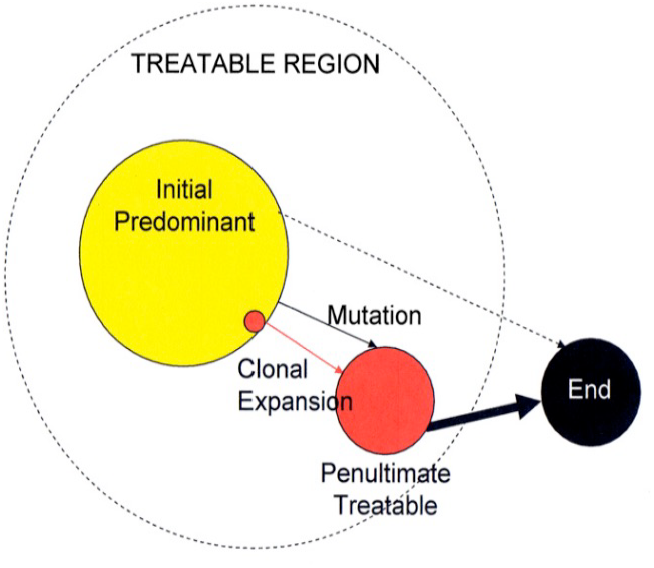
Graphical representation of phenotypic states in genetic space, illustrating the principles underlying DPM. The large dashed circle represents the genetic space that is treatable by available therapies. The three solid circles represent genetic states that correspond to phenotypic resistance states. The large yellow circle is the predominant state of the majority of cells at initial diagnosis (“initial-predominant state”). The black circle, outside the treatable space, is a lethal incurable state (“end state”). The red circles within the treatable space are “penultimate treatable states,” in that they are genetically close to the incurable (and multi-drug resistant) end state. Their smaller size indicates that at initial diagnosis, they had fewer cells, and perhaps were even undetectable. The medium sized red circle can arise by clonal expansion from the smaller red circle or by acquired mutation from the yellow circle and subsequent clonal expansion. The threat from the initial-predominant state is significant (yellow) because it contains the largest tumor burden. However, the threat from the penultimate-treatable state may be greater (red) because of its genetic proximity to the end state. Black arrows represent genetic transitions to resistance, the dashed black arrow indicates that several steps are required, and solid arrows represent single-step transitions. The thick arrow denotes a faster transition, either because there are multiple parallel genetic pathways to the phenotype or because the starting state is a hypermutator. Adapted from Ref [10] (Beckman, Schemmann, Yeang, PNAS 2012)

DPM changes therapy very frequently and plans ahead for potential future evolution of rare hypermutator subclones and others using an adaptive evolutionary model to predict the optimal treatment regimen within a series of short time windows (as short as 6 weeks), while taking into account predicted future events. DPM is pro-active compared to other approaches, which ‘react’ to new molecular findings either at relapse or progression or when detected earlier by liquid biopsy.

In a simulation of 3 million virtual patients, where each patient represented a unique set of input parameters derived from literature and clinical experience (cell number of each drug sensitivity phenotype, net growth rates, drug sensitivities, and net mutation and/or phenotypic evolution rates), DPM more than doubled median survival and markedly increased long-term survival and cure rates over CPM. This benefit is driven by approximately 30% of the virtual patients [7].

DPM provides optimal personalized treatment sequences for each virtual patient, divided into six-week treatment intervals called “moves”. We have further noted, in spite of its long-term benefits, that for most virtual patients, the benefit of DPM is achievable in the first two moves, greatly reducing the cost and invasiveness of measuring needed input parameters in patients [10]. This finding further enables us to identify a subset of virtual patients who are predicted to benefit from DPM [10]. In short, if the first two moves recommended by DPM are predicted to be identical to those from CPM, the patient is unlikely to benefit from DPM relative to CPM. This multicomponent “evolutionary classifier” is unusual in that it focuses on estimated growth rates, mutation rates, and drug sensitivities, i.e. dynamic properties of the cancer, rather than static molecular measurements [11-15].

The above advances make future clinical evaluation of DPM increasingly feasible. It thus becomes of interest to explore study designs to test DPM efficiently and accurately. Of note, classical study designs are meant to test specific drugs, whereas here we are attempting to test a strategy for personalized adaptive sequencing of the same drugs, including when to provide simultaneous combination therapy (possibly at reduced dose as often required for safety), when to provide pulses of high dose monotherapy, and when to change therapy completely.

Biomarker-strategy designs [11,13], Figure 2a, are utilised to evaluate the effectiveness of a new strategy over the existing control strategy. In such a design, patients are randomly assigned to the experimental treatment arm (that uses the biomarker to direct therapy) or to a control arm that does not. This then facilitates comparison of a new treatment strategy arm based on the biomarker to the standard-approach arm that does not consider the biomarker. However, a significant number of patients will receive the same treatment on either arm i.e. patients in the control strategy as well as patients who are biomarker negative in the biomarker-directed arm. This dilutes the between-arms treatment effect leading to loss in statistical power and increased sample size.

**Figure 2.**
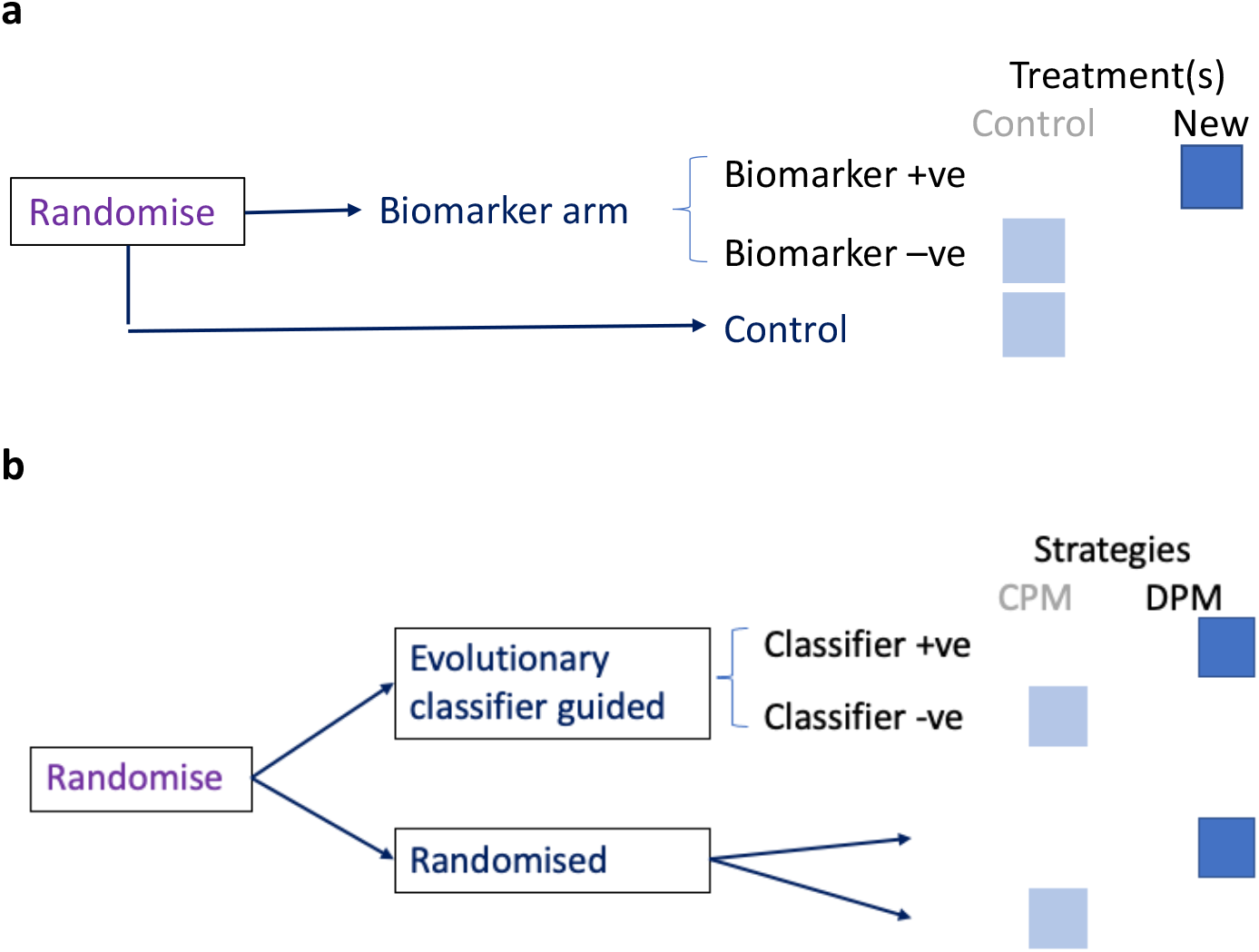
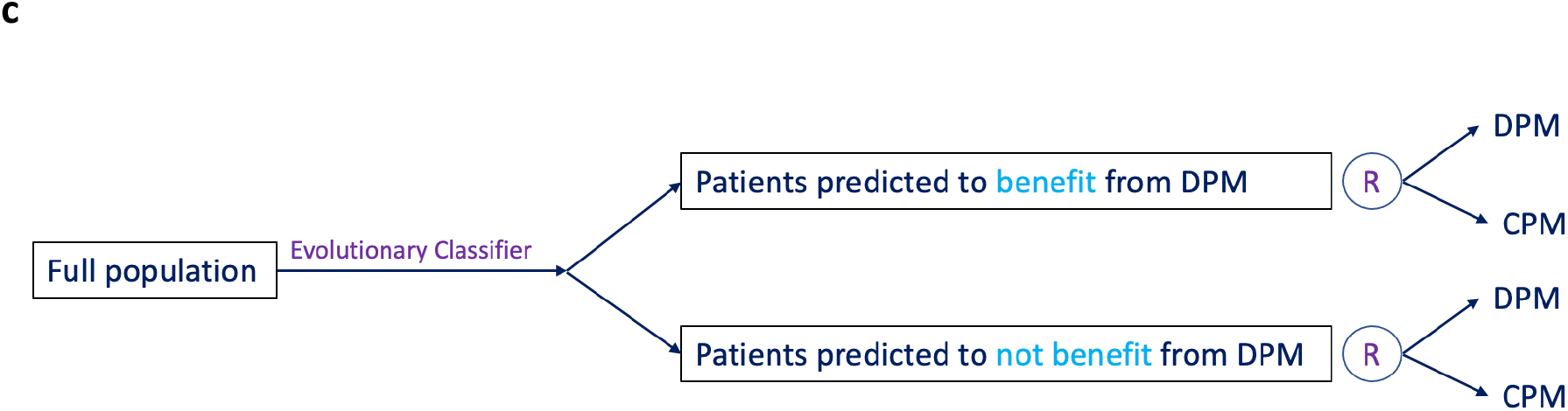
Strategy and Strategy-Stratified clinical trial designs. **a**. conventional biomarker-strategy design where patients are randomly assigned to experimental treatment arm (that uses the biomarker to direct therapy) or to a control arm that does not. **b**. candidate strategy design for DPM based on **a**; no insight into how much each cohort benefited from DPM. **c**. Proof-of-concept strategy-stratified design to evaluate DPM strategy effectiveness against CPM for each cohort.

If we were to adapt a simple strategy design for DPM using the evolutionary classifier as a biomarker, then patients in the evolutionary classifier cohort would be assigned DPM if and only if they were classifier positive, whereas patients in the non-evolutionary classifier cohort would be randomly assigned to DPM or CPM as in Figure 2b. However, such a design will yield no insight into how much each of the cohorts defined by the evolutionary classifier benefitted from DPM.

We, therefore, propose a bespoke strategy-stratified proof-of-concept clinical trial design, Figure 2c, that evaluates superiority of DPM strategy over CPM in patients predicted to benefit from DPM. Given that the role of a “biomarker” is now played by our Evolutionary Classifier, in the new design, each patient is assessed by the classifier that predicts whether or not they will benefit from DPM, which forms the stratification part of the design. The strata or cohort of patients predicted to benefit from DPM are then randomly assigned DPM or CPM strategies. The same is true for the cohort of patients predicted to not benefit from DPM. The design evaluates the benefit of DPM in both classifier positive and negative patients as well as testing the classifier.

However, a key challenge is that the evaluation of DPM within a clinical trial could be confounded by *incorrect estimates of drug sensitivity values*, which are input into a computer model of DPM by the doctor at the beginning of the study. In our model, we use at least two hypothetical drugs or drug combinations, with drugs (or combinations) 2 and above being considered late line therapies. Typically, cancer treatment proceeds in “lines of therapy”, progressing from first line therapy to later lines at each relapse. However, DPM does not wait until frank relapse and proactively calls for late line drugs earlier to minimize the risk of future relapse. Thus, in this scenario, we would lack clinical information on the performance of the later line drug when used as a first line therapy. This lack of data may be a challenge for a clinical study design. In the end, results may be confounded by the possibility that the later line drug is more effective in the first line than expected. Consequently, DPM should not solely be credited with this benefit, as hypothetically moving the drug forward in therapy would benefit patients, regardless of the use of DPM. This phenomenon could also work in the opposite way, with the late line drug performing worse than expected in first line therapy. Moreover, mis-specified drug sensitivity values may generally impair the ability of DPM as a decision tool.

To evaluate these concerns, we have performed “mis-specification simulations”. In these simulations, the doctor is given information that may be incorrect or noisy and inputs this information into a DPM based decision tool, making the best decisions possible under the circumstances, while the cancer evolves according to the correct values. Details are given in online methods and supplementary materials. In addition to meeting the usual performance metrics, clinical trial designs for testing DPM and other EGPMs need to be evaluated for robustness to incomplete or incorrect information, as they generally require more information for optimal performance, and it may be difficult to obtain this information in the clinic.

Our results confirm that our trial design has high power, controls false positive rates, and while affected to a degree by mis-specified values of drug sensitivity to drug 2, is surprisingly robust.

## Results

### Virtual Protocol

We utilized the simulated dataset of nearly 3 million virtual patients from Ref [7]. The patient population was divided into two distinct cohorts: patients expected to benefit from DPM, and patients not expected to benefit. If a patient’s first two moves were projected to be the same in DPM and CPM, they were put in the cohort *not* expected to benefit, while if either of their first two moves in DPM were different than those of CPM, they were put into the cohort expected to benefit. The benefit and non-benefit cohorts were then randomized into two groups: patients who would receive CPM and patients who would receive DPM. This protocol was created in order to test how the two cohorts overall benefited from DPM. In order to conduct the log rank test, the cox proportional hazards model was fitted, and Kaplan-Meier curves were plotted for this data. Datasets containing the CPM and DPM arms were combined, resulting in two overall datasets; one for each cohort. These data gave us the hazard ratio values, which compare the probability of survival in the treatment group (DPM) to the probability of survival in the control group (CPM). Figure 3 displays the survival curves for DPM and CPM patients in the benefit cohort and patients in the non-benefit cohort, respectively, in the absence of mis-specification. Note there is slight benefit in the non-benefit cohort too, i.e. a hazard ratio of 0.87, but in the benefit cohort the hazard ratio is 0.46.

**Figure 3.**
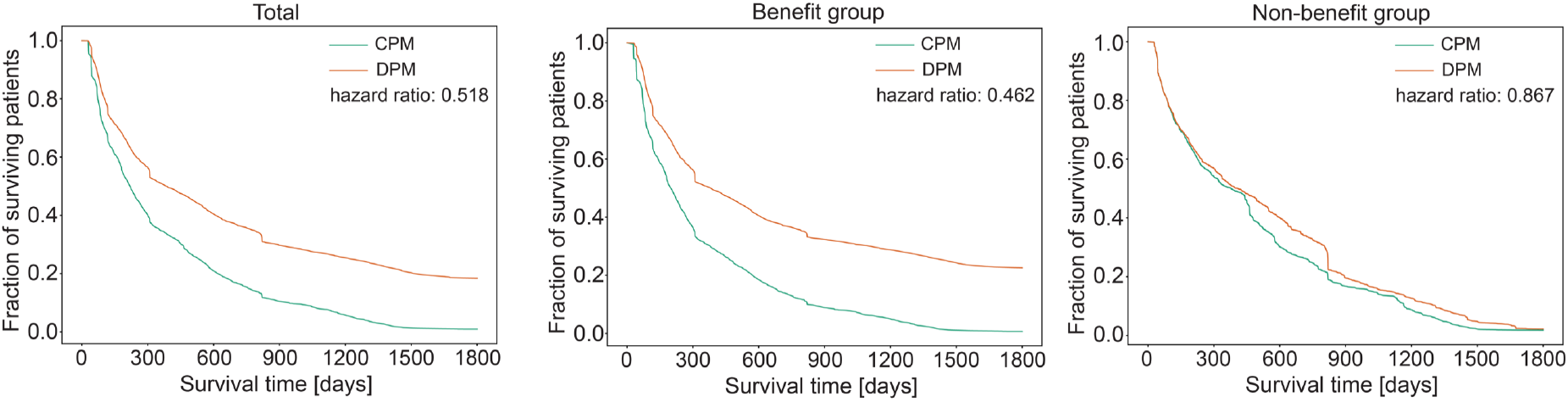
Kaplan-Meier curves comparing entire population. Three curves compare the total, benefit and non benefit chohort of CPM and DPM survival times. The benefit cohort benefits significanlty compared to the non-benefit cohort.

### Sample Size

Next, we determine the sample size required for a clinical trial with an overall survival endpoint. Note that since the full dataset of 3 million patients shows that the benefit cohort benefits significantly compared to the non-benefit cohort, we power the study on the Benefit cohort only. To model a real clinical trial, we consider an accrual period of 2 years and a follow-up period of 2 years for a study with 1:1 randomisation. Using functions in R, the number of events necessary for a study with a hazard ratio of 0.7, a type I error of 5%, and a type II error of 20% would be 194. Taking into account the probability of those events happening and factoring in a 15% dropout rate, 250 patients will be required from the Benefit cohort (125 each in DPM and CPM arms) with a power of 80% to detect the treatment effect with a hazard ratio of 0.7, i.e. 30% improvement in overall survival from DPM. Our hazard ratio assumption is more conservative than would result from the simulation (0.46), noting that the mathematical model does not account for all the complexities of cancer. A hazard ratio of 0.7 is considered to be clinically significant.

### Benefit Analysis

250 patients were randomly selected from our virtual patient pool and randomized equally into a CPM arm and a DPM arm. This was conducted twice, once for the Benefit cohort and once for the Non-Benefit cohort. After conducting 10,000 simulations of random subsets of 250 patients for each cohort, mean hazard ratios were calculated. This was done in order to compare the probabilities of survival in DPM with that in CPM.

For the Benefit cohort, the mean hazard ratio was 0.460, which compared to the value that our study was powered on (0.7), shows highly significant benefit, in line with prior results [7]. The mean hazard ratio from the benefit cohort was much lower than the mean hazard ratio of the non-benefit cohort, and therefore, we judged that the evolutionary classifier was effective in this simulated clinical trial setting as previously shown for large scale simulations [10].

### False negative rate with and without mis-specification

We defined a false negative when the cohort expected to benefit based on our large prior simulations did not benefit in the clinical trial simulation. Since our study was powered on a higher hazard ratio, the false negative rate among 10,000 trial simulations was 0% in the absence of mis-specification, as shown in Figure 4a.

**Figure 4.**
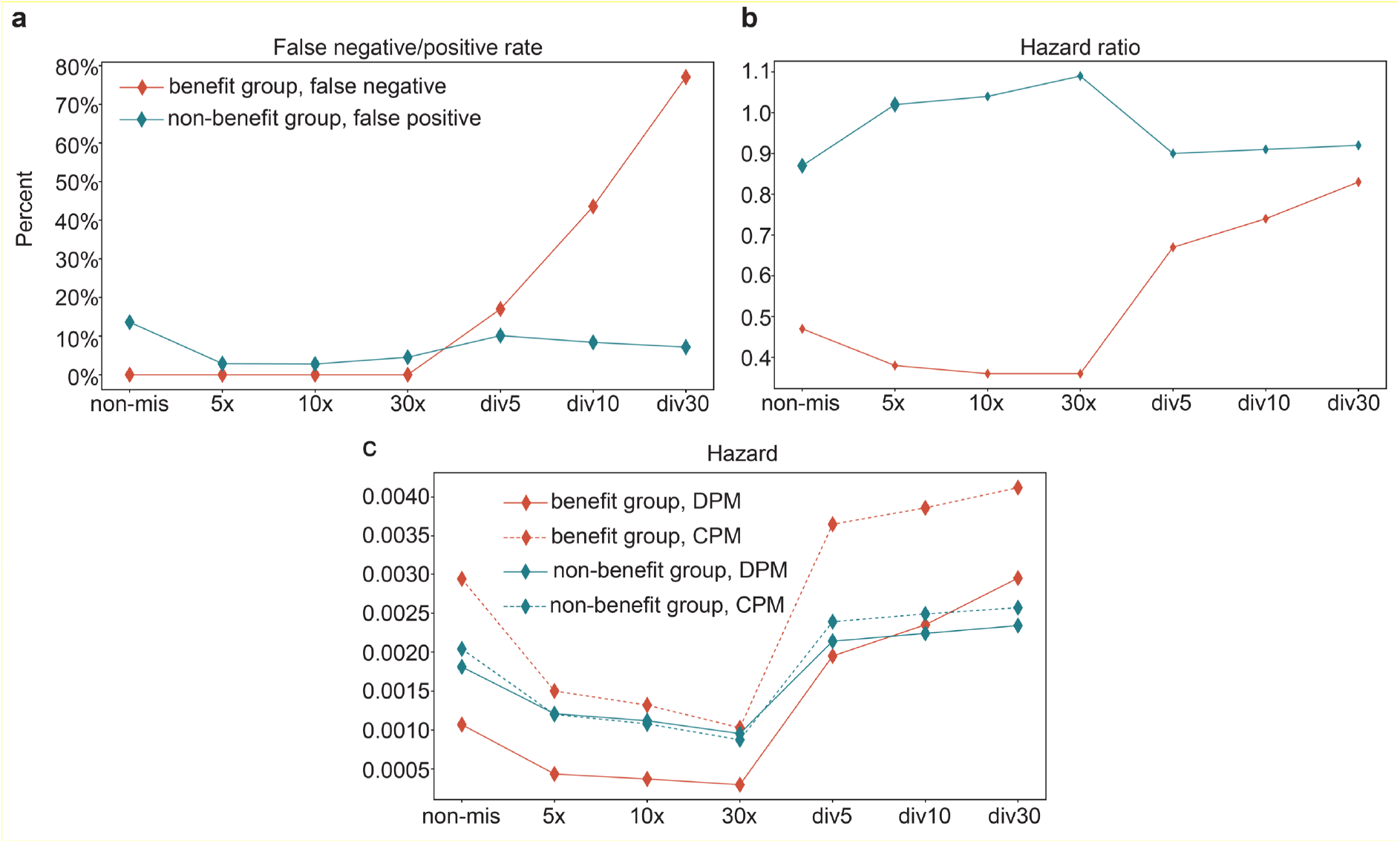
Comparison of false negative/positive rates, hazard ratios and hazards with and without mis-specification for benefit and non-benefit groups. **a**, Changes in the false negative rate for the benefit group and the false positive rate for the non-benefit group with and without various mis-specifications of drug 2 efficacy. **b**, Similar to **a**, but for the changes in hazard ratios for both benefit and non-benefit groups. **c**, Similar to **a**, but for the changes in the hazards of CPM and DPM separately for both benefit and non-benefit groups.

When the true sensitivity to drug 2 was higher than expected (5X, 10X and 30X), the false positive rate continued to be zero as expected.

When the true sensitivity to drug 2 was lower than expected (div 5, div 10, and div 30), false negatives for DPM began to increase, as expected. Nonetheless, due to the conservative powering assumption, the false negative rate was less than 20% at div 5 (sensitivity to drug 2 1/5 of that expected), corresponding to greater than 80% power in that scenario. If drug 2 was still weaker, the false negative rate concerning DPM rose to higher levels, indicating that the benefit of DPM cannot be seen if drug 2 underperforms by 10-fold or more relative to expectations.

### False positive rate with and without mis-specification

We defined a false positive when the cohort expected not to benefit based on our large prior simulations benefitted in the clinical trial simulation. The study was designed to have a 5% false positive rate; however, it is important to note that this rate is normally defined when compared to an absolute negative, which would have had a hazard ratio of 1.0. Since the non-benefit group had some benefit (hazard ratio 0.872) some of what we defined as false positives were likely true positives. In fact, with a hazard ratio of 0.872 and 125 patients per arm, the true positive rate could be as high as 16%. That is, a study of this size has 16% power to show that a true hazard ratio of 0.872 is better than a hazard ratio of 1. To further clarify, the non-benefit population should have had a hazard ratio of 1, which would mean they did not benefit at all. The fact that they demonstrated some benefit (hazard ratio of 0.872) means that the false positive rate should be higher than a non-benefit population that demonstrated absolutely no benefit.

For the Non-benefit population, the mean hazard ratio was 0.872, with a false positive rate of 12.4% in the absence of mis-specification, Figure 4a, indicating a very low rate of false positives.

Our original hypothesis was that if sensitivity to drug 2 was greater than expected (5X, 10X, or 30X), this would result in an increased rate of false positives, falsely concluding DPM had merit in the non-benefit population. However, this was not observed. This was apparently because CPM also benefitted from increased sensitivity to drug 2, which allowed it to more effectively salvage the patients even though it waited until frank relapse to utilize drug 2 (Figure 4c). When drug 2 was more effective than expected, absolute hazards for both DPM and CPM decreased in the non-benefit group, but their ratio actually increased slightly, resulting in a lower “false positive” rate, which in fact may have represented fewer true positives, contrary to our hypothesis.

### Benefit of DPM as a function of mis-specification

As shown in the Figure 4b, the benefit in the benefit group for DPM relative to CPM increased slightly when drug was more potent than expected, but decreased significantly when drug 2 was less potent than expected, especially if the discrepancy was more than 5-fold. As discussed above, the small effect on DPM benefit when drug 2 was more potent was unexpected, and apparently due to a nearly equal increase in the efficacy of CPM (Figure 4c). In the non-benefit group, the slight relative benefit of DPM lessened when drug 2 was more potent than expected, apparently due to a greater decrease in the hazard ratio for CPM (Figure 4c).

### Utilization of drug 2 in DPM compared to CPM as a function of mis-specification

We utilized 3 metrics for drug 2 utilization: 1. Average move of first introduction of drug 2; 2. Average dose intensity per move of drug 2 throughout the treatment course, and 3. Average move number of drug 2 utilization weighted by dose intensity. This is a measure of how late drug 2 is given. Details are given in online methods.

As shown in Figures 5a and 5c, and in accord with our hypothesis, in the benefit group DPM initially calls for drug 2 earlier and uses drug earlier on average compared to CPM. This is relatively unaffected by the actual drug 2 potency, which is also not surprising since the doctor is given the same information in every case, it is the ground truth that is varied.

**Figure 5.**
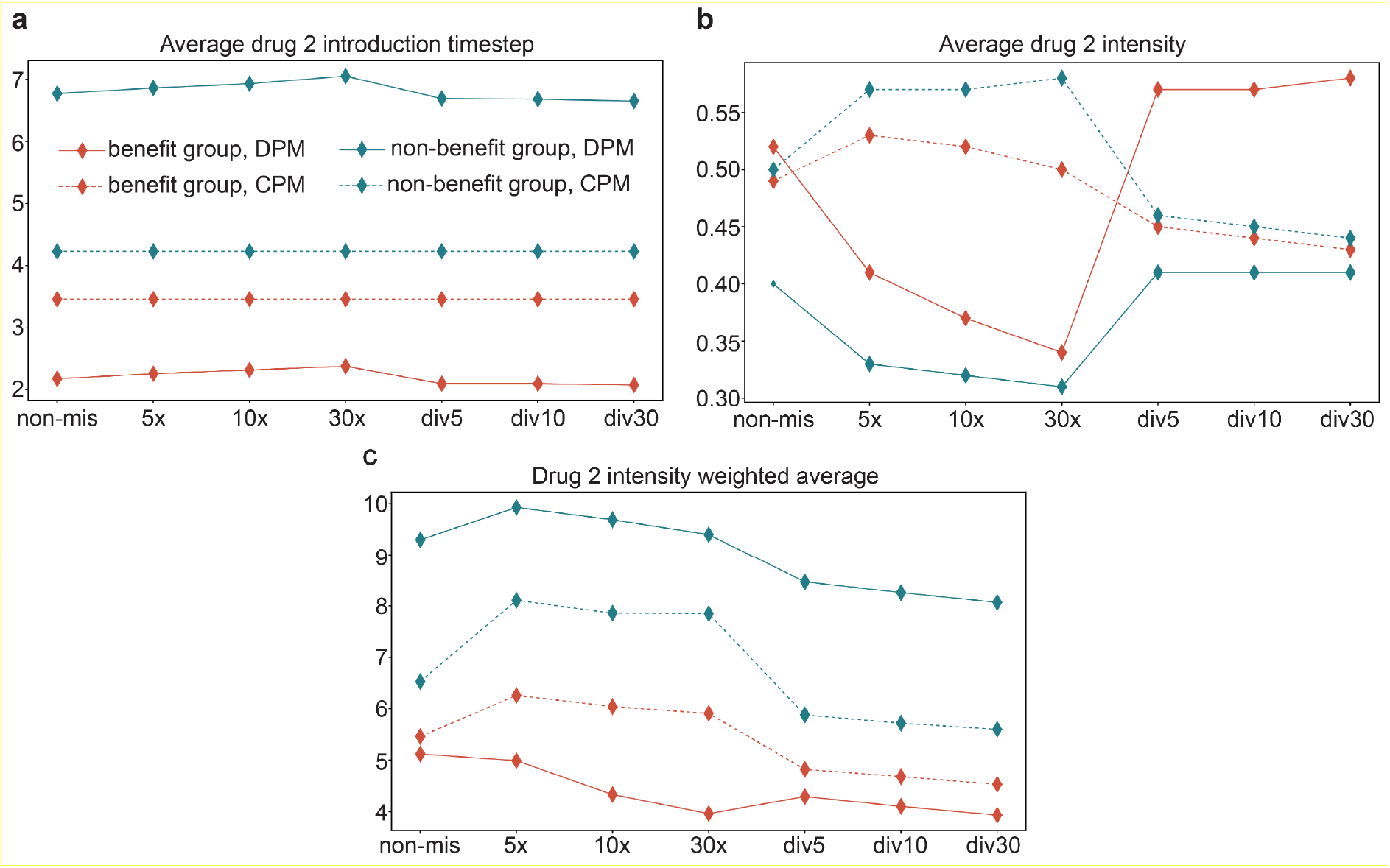
Comparison of metrics evaluating the use of drug 2 across scenarios with and without mis-specifications for benefit and non-benefit groups. **a**, Changes in average drug 2 first use timestep for CPM and DPM in both the benefit and non-benefit groups. **b**, Similar to **a**, but for changes in average drug 2 intensity. **c**, Similar to **a**, but for the changes in drug 2 intensity weighted average timestep of use. See online methods for further definition of metrics.

In contrast, in the non-benefit group it is CPM that initially calls for drug 2 earlier and uses it earlier on average compared to DPM, and again this is relatively unaffected by mis-specification. The reason for this trend requires further investigation. We note that by convention drug 1 is the usual first line drug, and the non-benefit group is defined by identical recommendations by DPM and CPM for the first 2 moves.

The average dose intensity for drug 2 is similar for DPM and CPM in the benefit group in the absence of mis-specification (Figure 5b). Although DPM calls for drug 2 earlier in the benefit group, elimination of the drug 1 resistant subclone early prevents relapse and later use of drug 2. Average dose intensity for drug 2 is slightly lower for DPM than CPM in the non-benefit group, a phenomenon that requires further investigation.

Average dose intensity for drug 2 declines significantly for DPM in both cohorts, if drug 2 is more potent than expected, presumably reflecting the more rapid elimination of drug 1 resistant subclones when drug 2 is more potent than expected.

## Discussion

The high risk of drug resistance due to pre-existing intratumoral genetic heterogeneity as well as subsequent acquired mutations limits the benefit of current precision oncology. Evolutionary guided precision medicine (EGPM) is a family of approaches that holds promise for improving overall survival by delaying the acquisition of drug resistance. One form of EGPM, adaptive therapy, has achieved promising results in prostate cancer in an unrandomized pilot study [16]. However, as EGPM is clinically tested beyond pilot studies, there will be a need to develop randomized proof-of-concept and confirmatory study designs that test the overall strategies without confounding by unanticipated and potentially unmeasured variables related to a specific application example. For example, as we have outlined, an EGPM that uses a particular drug more will appear more or less beneficial if that drug is more or less effective than expected in general. Such confounding can interfere with generalizability of conclusions concerning the merit (or lack thereof) of an EGPM.

In this study, we have simulated a randomized “stratified strategy” proof-of-concept trial design for evaluation of dynamic precision medicine. The design is unusual in that while stratified study designs are common and “strategy” designs are common, the application of stratification to “strategy” designs is less common. The stratification is also not by a conventional biomarker or prognostic characteristic, but by a unique multicomponent classifier featuring dynamic evolutionary properties rather than static molecular measurements. The classifier has been shown to predict DPM benefit in larger simulations [10].

We simulated the study design of 500 total patients (250 per stratum, randomized to equally between DPM and CPM) 10,000 times by randomly sampling a previously constructed database of approximately 3 million virtual patients representing varied clinical presentations [7]. We find that the design has high power and a low false positive rate, reproduces the hazard ratio expected from simulations, and also reproduces results from larger simulations on the evolutionary classifier.

We further tested the robustness of the design to mis-specification of the potency of “drug 2”, a late line drug that might be called earlier by DPM than CPM, since data on the clinical performance of late line drugs when given earlier would be limited. Contrary to our hypothesis, the false positive rate (falsely assigning merit to DPM in the non-benefit population) remained low, even when drug 2 was as much as 30 times more potent than expected, because a more potent drug 2 benefitted DPM and CPM equally, a highly reassuring finding since false positives would lead to expensive confirmatory testing of DPM. Our conservative powering strategy also maintained power and avoided false negatives when drug 2 was 5-fold less potent than expected, but power rapidly declined if drug 2 was still weaker than that. This should be kept in mind in interpreting negative results from a single EGPM study.

The findings in the benefit cohort with respect to mis-specifications are well explained by the different patterns of usage of drug 2, including DPM calling for drug 2 earlier but also terminating its use earlier upon elimination of drug 1 resistant subclones. Some of the trends observed in the non-benefit cohort require further investigation.

Five hundred patients are rather large for a proof-of-concept study, and the size might be reduced by interim futility analyses in both cohorts in a future iteration of the design. However, it is well to remember that these proof-of-concept studies are testing a strategy which, depending on its estimated merit at proof-of-concept, might or might not be very broadly applied.

In summary, a novel clinical study design for DPM has shown good statistical properties and overall robustness to mis-specification of drug 2 sensitivity, and may be a useful randomized proof-of-concept design for EGPM in general.

## Online Methods

### Identification of Benefit and Non-Benefit Cohorts

The EC developed in [10] matched patients to therapies not just based on static molecular properties but also on dynamic properties such as growth and mutation rates that may predict future state. It was also shown that the 2 move (point of therapy adaptation for a 45-day cycle per move) window intervention provided benefit similar to the full 40 moves of DPM. Recall that Benefit was defined by an overall increase in survival by at least 25%, and at least 60 days when using the DPM strategy.

We now define the Benefit and Non-Benefit cohorts. Specifically, if either of the first two projected moves recommended to a patient by DPM were different than that of CPM, prior simulations [10] have shown that these virtual patients receiving DPM would indeed benefit. Thus, this virtual population is known to be a *true positive population* (a population that benefits from DPM) in simulations, or simply the Benefit cohort. Additionally, this population can also be used to determine the false negative rate of any clinical trial design.

To clarify, if the first two projected moves recommended by DPM are identical to CPM’s first two projected moves, the virtual patient would *generally* not benefit from receiving DPM [10]. Thus, this population of virtual patients is an *approximate true negative population* in simulations, or simply the Non-Benefit cohort.

Let us now look at false negatives and false positives. The Benefit cohort can be used to determine the false negative rate; specifically, a false negative would be a population that we expect to benefit from DPM, but in a simulated clinical trial, does not. Likewise, the Non-Benefit cohort can be used to determine the false positive rate of a proposed clinical trial design within limits, given that the Non-Benefit population is an approximate true negative. Specifically, a false positive would be those who we do not expect to benefit from DPM, but in simulation, do. In our study, since patients within both the true positive and approximate true negative cohorts are randomized into either DPM or CPM strategies, this provides insight into the false negative and false positive rates in the stratified-strategy design under different conditions.

### Drug 2 Sensitivities

#### Model simulation

Simulation studies are carried out to evaluate the effectiveness of different treatment methods by different virtual patient settings. There are a total of 4 cell states in the mathematical model: *S, R*_1_, *R*_2_ and *R*_12_ cells, which are sensitive to both drugs, resistant to drug 1 and sensitive to drug 2, resistant to drug 2 and sensitive to drug 1 and resistant to both drugs, respectively. The initial total cell population is 5 × 10^9^, roughly equivalent to a 5-cm^3^ lesion. The initial cell number for *R*_1_ and *R*_2_ cells are set by parameterS 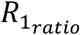 and 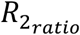. The timestep for predicting responses in DPM is 45 days, which is to mimic 6 weeks that elapse between evaluations on clinical studies. The dosage of two drugs, *σ*1 and *σ*2 respectively, was chosen from among 0, 0.5 and 1 with the constraint that the sum of the dosages of the two drugs equal to 1, to simulate needed dose reduction in combination. In real applications, needed dose reductions in combination would be governed by combination Phase 1 study results. So, there are 3 possible dosage selections of the two drugs 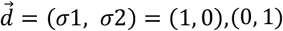. Simulation stopped for mortality if the total cell number of virtual patient exceeded 10^13^, or the virtual patient is cured (cell number of each cell state smaller than 1). The number of 10^13^ is intended to represent the sum of total cell number in many metastatic lesions as most patients succumb to metastasis. Cure is defined as total elimination of cells.

#### Treatment methods

CPM (Current Precision Medicine): Initially, treat the virtual patient with the most effective drug on the predominant cell state selected by molecular characterization. A nadir is a local minimum of the total cell number among the time-series profile where the current treatment is maintained. At *t* = 0, nadir is equal to the initial total population which is 5 ×10^9^. Change the drug if either one of the following events happens: (i) the total cell number reaches twice the nadir or (ii) the total cell number reemerges from a level below the detection threshold (10^9^). If either (i) or (ii) happens and another drug has not been used, switch to another drug which means that each drug only be used once. Update the nadir at each timestep.

DPM (Dynamic Precision Medicine): Minimize the number of incurable cells (*R*_12_ cells) unless there is an urgent clinical need for cytoreduction. In the simulation, this is set at a threshold cell number, but in practice it would vary greatly depending on the clinical circumstances and is left to the judgement of the clinician, for whom DPM is provided only as a decision tool. The discussion below assumes an arbitrary threshold of 10^11^ cells. At each timestep, select the 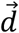 from (1, 0), (0, 1) and (0.5, 0.5) that gives the minimum predicted cell number of *R*_12_ cells if the total cell number does not exceed 10^11^. *R*_12_ cells are resistant to both drugs, and therefore incurable. By preventing the formation of *R*_12_ cells, the possibility for long-term survival and/or cured is maintained. If the total cell number exceeds 10^11^, select the 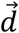 from (1, 0), (0, 1) and (0.5, 0.5) that gives the minimum predicted total cell number.

#### Mis-specification of Drug 2 effect

In order to test the influence of the uncertainty of drug 2 efficacy on CPM and DPM treatment strategy, mis-specifications of drug 2 are introduced. In the model simulation, the true effect of drug 2 ranged from 1/30th to 30 times the effect of drug 2 used in the doctor’s decision. At each timestep, doctors will decide the drugs dose according to a strategy and *S*_2,1_ is used, which is the sensitivity of *S* cells to drug 2 relative to drug 1. After that the model simulation is performed under the selected drug dose but based on true *S*_2,1_ which termed as 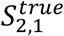. The possible values of 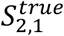 are *S*_2,1_/30 (div30), *S*_2,1_/10 (div10), *S*_2,1_/5 (div5), *S*_2,1_ × 5 (5x), *S*_2,1_ × 10 (10x), *S*_2,1_ × 30 (30x). The model simulation under the decided dose is performed based on the true drug 2 effect. This process is continued to the end of the total time considered (5 years) or the simulation stopping criteria are met.

#### Benefit group and non-benefit group

Benefit group and non-benefit group are defined without the mis-specification situation (total number of virtual patients: 3,083,233; number of virtual patients in benefit group: 2,450,211, number of virtual patients in non-benefit group: 633,022). Benefit group is classified by evolutionary classifier, which are the virtual patients whose drug selections at one or more of the first two timestep are different between CPM and DPM. Vice versa, the non-benefit group are the virtual patients whose drug selections at the first two timestep are identical.

#### Definitions for Analysis

Average drug 2 introduction move is defined as 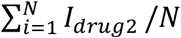, which is the averaged move index where drug 2 was first used. *I*_*drug*2_ is the index of the move where drug 2 was first used. *N* is the total number of virtual patients in the interested group and *i* is the ith virtual patients. As the maximum number of moves is 40, each timestep is 45 days and simulation period are 1800 days, the possible value of *I*_*drug*2_ can be from 1 to 40. If drug 2 has never been used for a specific patient, *I*_*drug*2_ is set to the maximum number of moves which is 40.

Averaged drug 2 intensity is defined as 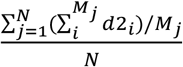, which is the averaged mean drug 2 dose intensity over the virtual patients of interest. *d*2_*i*_ is the drug 2 dose at *i*th timestep for *j*th virtual patient, with the possible value 0, 0.5 and 1. *M*_*j*_ is the total number of moves for *j*th virtual patient, with the possible value from 1 to 40. *N* is the total number of virtual patients in the group of interest.

Drug 2 intensity weighted average is defined as 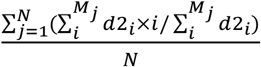, which is the averaged time of utilization of drug 2 weighted by dose intensity over the virtual patients of interest. *d*2_*i*_ is the drug 2 dose at *i*th timestep for *j*th virtual patient. *M*_*j*_ is the total number of moves for *j*th virtual patient, with the possible value from 1 to 40. *N* is the total number of virtual patients in the interested group.

Hazard ratios are calculated by the Cox proportional hazard model and hazard ratios are calculated by fitting exponential model. The confidence intervals in Supplementary Material are calculated by bootstrap resampling, in which case the 25^th^ and 975^th^ of 10,000 resampled with replacement mean values are the lower and upper confidence interval, respectively.

## Data Availability

All data produced in the present study are available upon reasonable request to the authors

## Acknowledgements

This work is supported by The Royal Society International Exchanges Award IES\R3\183092 to DP and RAB, and by Department of Defense (DoD) Breast Cancer Research Program Breakthrough Award W81XWH-20-1-0760 to RAB. RAB was partially funded by the National Institutes of Health Cancer Center Support Grant (CCSG) 5P30CA051008. CHY was partially funded by a grant (108-2118-M-001-001-MY2) from National Science & Technology Council in the Republic of China (Taiwan).

## Supplementary Data

### Model Parameters

There is total 9 parameters in the full model, which are:

1. 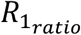, which is the ratio of the initial *R*_1_ cell number to the total. Possible values are 0, 10^−9^, 10^−7^, 10^−5^, 10^−3^, 10^−1^, and 0.9. *R*_1_ cell number varies from absent to five cells; from four 2-log (10^2^) increments to 10%; and the case where *R*_1_ cells dominate the population.
2. 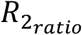, which is the ratio of the initial *R*_2_ cell number to the total. The values are the same as 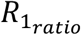.
3. *g*, which is the pre day net growth rate. Possible values are 0.001, 0.002642, 0.00698, 0.018439, 0.048714, 0.128696 and 0.34. The maximum value 0.34 represents a scenario assuming a 48-hour growth rate, 100% cells in cycle and zero death rate [1-3]. The minimum value 0.001 represents a slow-growing tumor that doubles in size in 2 years period. The low range is the range of growth rates that fit data from 228 patients [4]. The range was divided into equally spaced logarithmic intervals 0.972 between 0.001 and 0.34. A value of 0.055 corresponds to a tumor which doubles in size in the 6 weeks interval between evaluations on clinical studies.
4. *S*_1,g_, which is the sensitivity of *S* cells to drug 1 relative to the net growth rate *g*. Possible values are 0.00056, 0.005379, 0.051674, 0.496387, 4.76831, 45.804544 and 440. The minimum value 0.00056 is sufficient to cause a 25% increase in the progression-free survival (20% decrease in the instantaneous progression hazard) for the slowest growth rate 0.001 listed above, which corresponds to the minimum effect deemed of interest in clinical drug development. The maximum value 440 is to cause 4-log (10^4^) decrease in cell number in 3 months for tumor with the fastest growth rate 0.34 listed above. Between the extremes, the range was divided into equally spaced logarithmic intervals 2.262 between 0.00056 and 440.
5. *S*_2,1_, which is the sensitivity of *S* cells to drug 2 relatives to drug 1. Possible values are 0.0004, 0.001474, 0.005429, 0.02, 0.073681, 0.271442 and 1. We assumed that drug 1 is more effective than drug 2 against *S* cells so the highest possible value of this ratio is 1. The lowest possible value is the lowest sensitivity of interest divided by the highest sensitivity of interest. The range is divided into equally spaced logarithmic intervals 1.304 between 0.0004 and 1. *S*_2,g_, the sensitivity of *S* cells to drug 2 relatives to the net growth rate *g*, which will be *S*_2,1_ × *S*_1,g_.
6. *S*_*R*1,1_ which is the sensitivity of *R*_1_ cells to drug 1 over sensitivity of *S* cell to drug 1. Possible values are 0, 10^−5^, 9.564 × 10^−5^, 9.146 × 10^−4^, 8.747 × 10^−3^, 8.365 × 10^−2^ and 0.8. The values were selected arbitrarily to include 0 (complete resistance), 10^−5^ (considerable resistance) and 0.8 (minimal resistance). The range is divided into equally spaced logarithmic interval 2.258 between 10^−5^ and 0.8. *S*_*R*1,g_, the sensitivity of *R*_1_ cells to drug 1 relative to the net growth rate *g*, which will be *S*_*R*1,1_ × *S*_1,g_.
7. *S*_*R*2,2_, which is the sensitivity of *R*_2_ cell to drug 2 over sensitivity of *S* cell to drug 2. The values are identical to 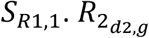, the sensitivity of *R*_2_ cells to drug 2 relatives to the net growth rate *g*, which will be *S*_*R*2,g_ × *S*_2,g_.
8. *T*_1_, which is the transition rate from *S* to *R*_1_ cells. Possible values are 10^−11^, 2.154 × 10^−10^, 4.642 × 10^−9^, 10^−7^, 2.154 × 10^−6^, 4.642 × 10^−5^ and 10^−3^. The transition rate is the sum of all possible transition rates over relevant loci that alter the sensitivity and resistance phenotype. The minimum value is the wild type of mutation rate estimated from human stem cells [5]. It is in accord with a mutation rate in genetically stable retinoblastoma, which is 2.2 × 10^−8^ in a full tumor, or less than 2.2 × 10^−10^ if 100 or more cell generations to develop the tumor are assumed [6]. The maximum transition rates assume a mutator mutation of 50000-fold increase in mutation rate [7] and many parallel pathways to acquire resistance. The range of 10^−11^ to 10^−3^ in accord with data from a series of patients with pancreatic cancer [4] and is divided into equally spaced logarithmic interval 3.07.
9. *T*_2_, which is the transition rate from *S* to *R*_2_ cells. The values are identical to *T*_1_. There is total 3,083,233 parameter configurations remained as our previous work used [8].

### Mathematical Model

The 4 different cell states are represented by a four-component vector, 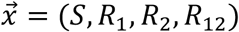. All cells have the same growth rate *g*. The cell death rates are set to 0 without treatment and the drugs work by increasing death rate. The model can be expressed by a differential equation:

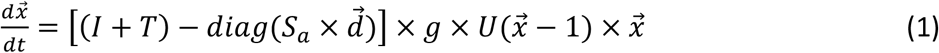

where *I* is a four-by-four identity matrix and *diag*(·) is an operator of placing vector components on the diagonal entries of a zero matrix. The growth rate *g* is same for all cell states. 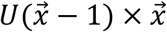 is the Heaviside step function that sets component values to 0 if they are less than 1, which means that *U*(*x*_*i*_ − 1) = 0 if *x*_*i*_ < 1 and *U*(*x*_*i*_ − 1) = 1 if *x*_*i*_ ≥ 1, *x*_*i*_ is the component in 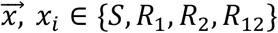. This term stipulates that fractional cell numbers (cell number less than 1) will not contribute to cell division. *T* is a four-by-four transition rate matrix corresponds to the transitions from all other cell types, which denoted as:

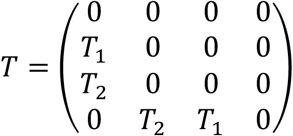

*T*(*i, j*) specifics the transition rate from cell state *j* to *i*. We assume that (i) transition rates from resistant to sensitive cell states are negligible, (ii) the transition rate of acquiring the resistance to one drug is independent of the resistance state to another drug, and (iii) transition rates of acquiring double resistance in one step are negligible.

*S*_*a*_ is a four-by-two matrix representing sensitivities relative to the net growth rate *g*, which denoted as:

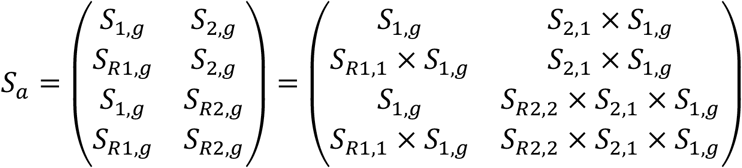

## Supplementary Figures and Tables

**Figure S1.**
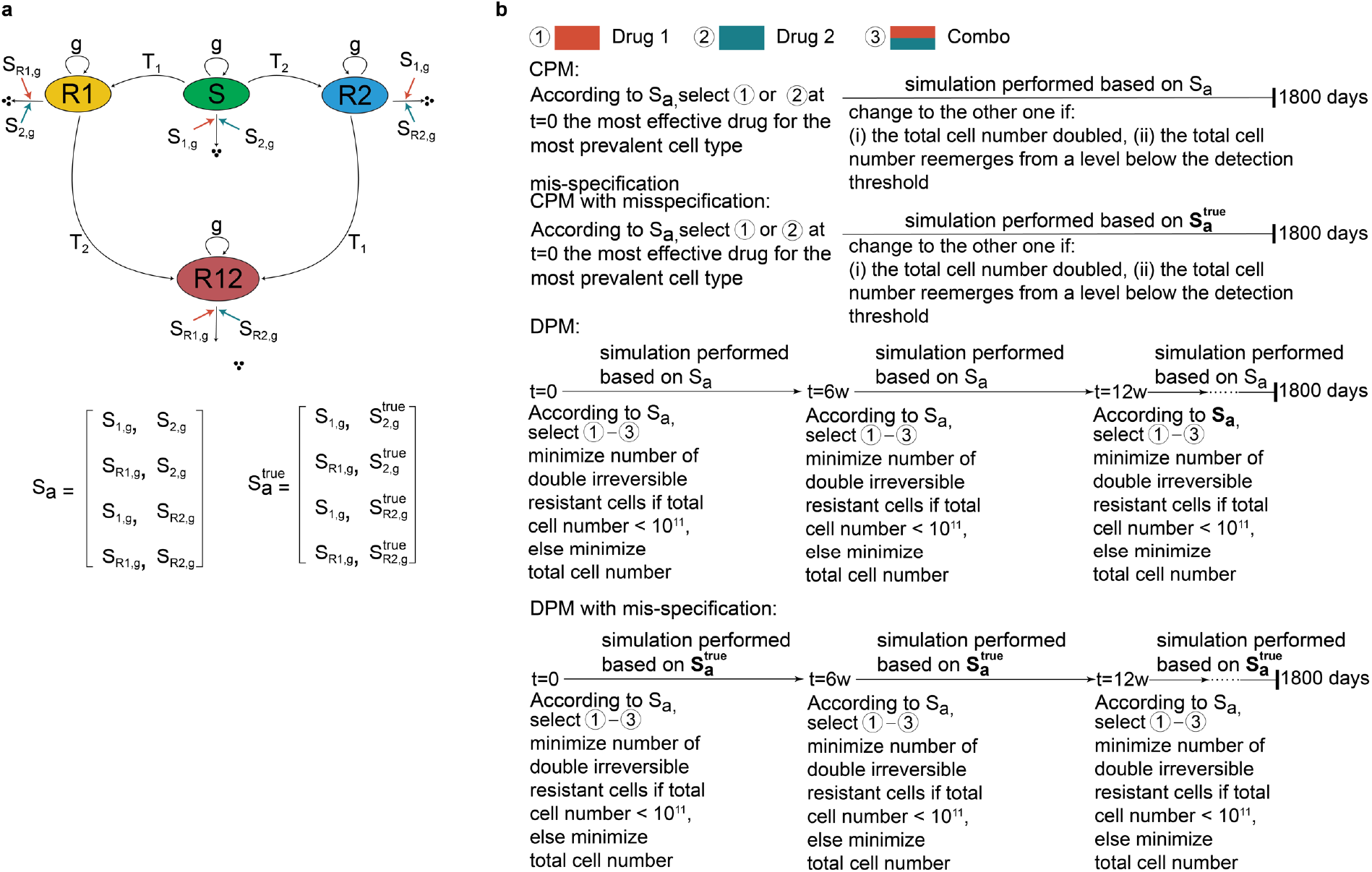
A schematic illustration of the mathematical model structures and the workflow for conducting simulations with and without mis-specification. **a**, The schematic illustrates the dynamics of four cell states and their transitions under the influence of drug 1 and drug 2, forming the structure for the simulation. The arrows represent self-replication, degradation and transitions between specific states. The *S*_*a*_ and 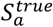 matrix are defined by their respective mis-specified and true individual drug sensitivities. **b**, Workflow for simulations with and without mis-specification in drug sensitivity. The total simulation duration is 1800 days, with a DPM strategy simulation timestep of 45 days, resulting in 39 steps.

**Figure S2.**
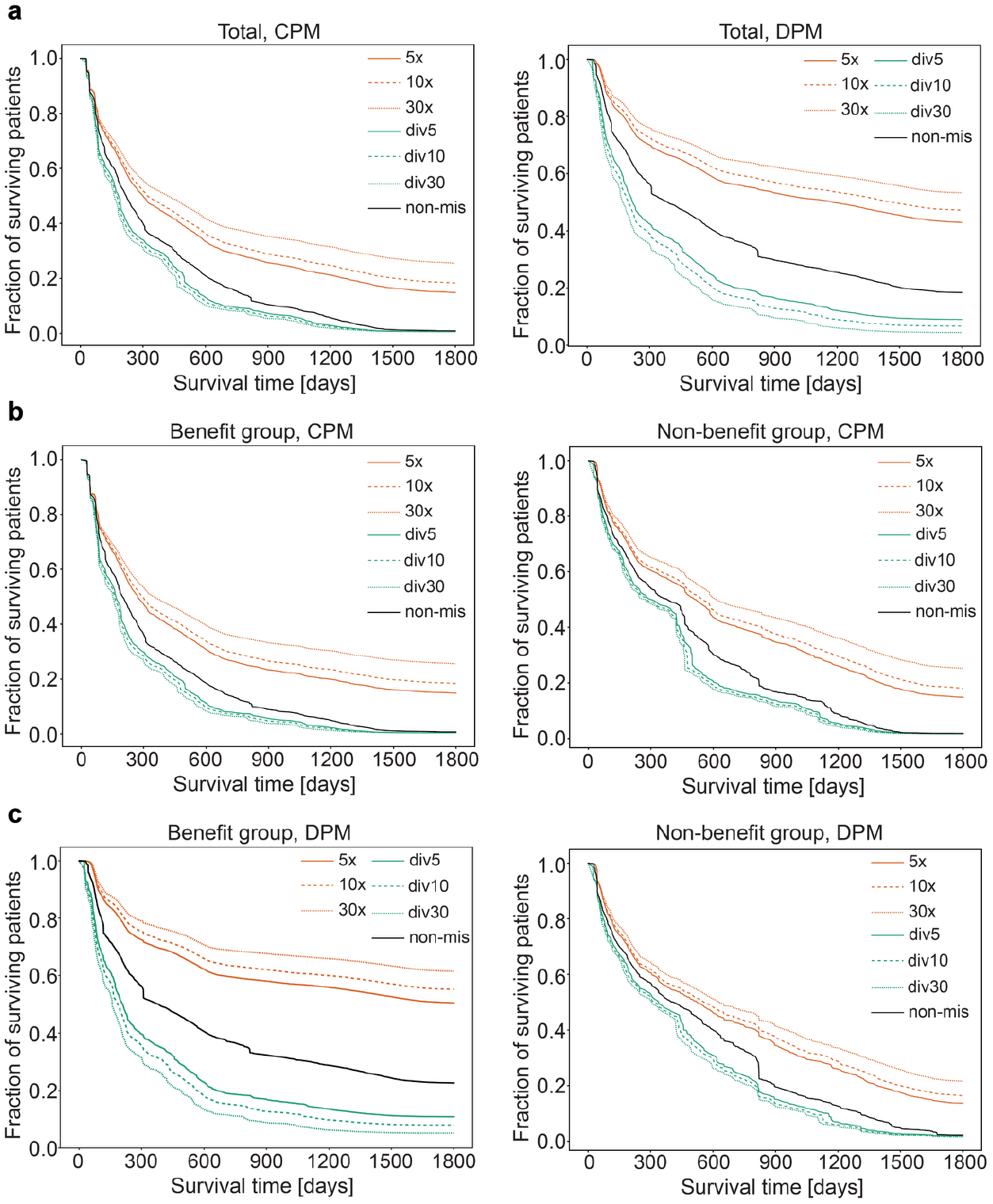
Kaplan-Meier curves illustrating the comparison between CPM and DPM treatment strategies under varying conditions, including the presence and absence of different mis-specifications across distinct groups. **a**, Kaplan-Meier curves showing CPM and DPM for the entire virtual patient group: CPM with mis-specification (left) and DPM with mis-specification (right). **b**, Similar to **a**, but for the benefit group. **c**, Similar to **a**, but for the non-benefit group.

**Table S1.**
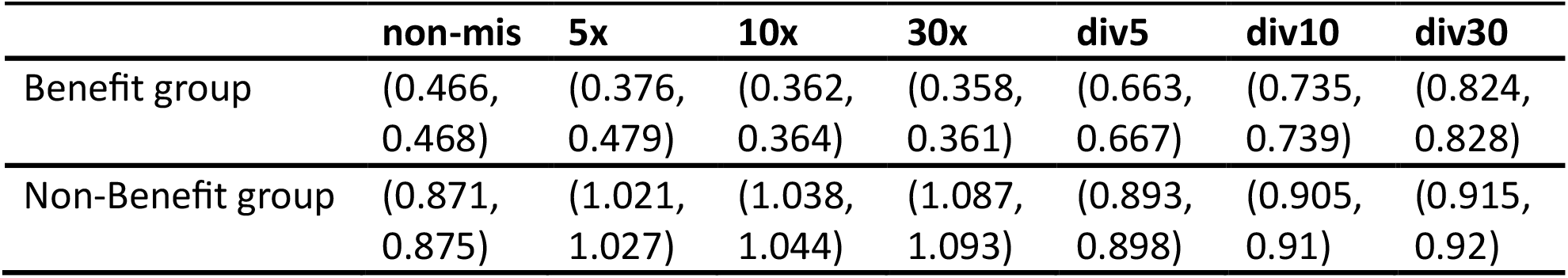
Confidence interval of hazard ratio in different groups with and without mis-specification at model simulation.

|  | non-mis | 5x | 10x | 30x | div5 | div10 | div30 |
| --- | --- | --- | --- | --- | --- | --- | --- |
| Benefit group | (0.466, 0.468) | (0.376, 0.479) | (0.362, 0.364) | (0.358, 0.361) | (0.663, 0.667) | (0.735, 0.739) | (0.824, 0.828) |
| Non-Benefit group | (0.871, 0.875) | (1.021, 1.027) | (1.038, 1.044) | (1.087, 1.093) | (0.893, 0.898) | (0.905, 0.91) | (0.915, 0.92) |

## References

1. Schmitt MW et al. Detection of ultra-rare mutations by next-generation sequencing. Proc Natl. Acad. Sci. 2012;109: 14508–14513.

2. Loeb LA, Kohrn BF, Loubet-Senear KJ, Dunn YJ., Ahn EH, O’Sullivan JN, Salk JJ, Bronner MP, Beckman RA. Extensive subclonal mutation diversity in human colorectal cancer and its significance. Proc Natl. Acad. Sci. 2019;116: 26863–26972.

3. Beckman RA. Neutral evolution of rare cancer mutations in the computer and the clinic. npj Systems Biology and Applications 2024. doi 10.1038/s41540-024-00436-3

4. Beckman RA, Loeb LA, Rare mutations in cancer drug resistance and implications for therapy. Clinical Pharmacology and Therapeutics 2020; 108:437–439.

5. Gatenby RA, Silva AS, Gillies RJ, Frieden BR. Adaptive therapy. Cancer Res. 2009; 69(11):4894–903. doi:10.1158/0008-5472.CAN-08-3658

6. Chmielecki J, Foo J, Oxnard GR, et al. Optimization of dosing for EGFR-mutant non-small cell lung cancer with evolutionary cancer modeling. Sci Transl Med. 2011; 3(90):90ra59. doi:10.1126/scitranslmed.3002356

7. Beckman RA, Schemmann GS, Yeang CH. Impact of genetic dynamics and single-cell heterogeneity on development of nonstandard personalized medicine strategies for cancer. Proc Natl Acad Sci 2012; 109(36):14586–91. doi:10.1073/pnas.1203559109

8. Leder K, Pitter K, LaPlant Q, et al. Mathematical modeling of PDGF-driven glioblastoma reveals optimized radiation dosing schedules. Cell. 2014; 156(3):603–616. doi:10.1016/j.cell.2013.12.029

9. Beckman RA, Loeb LA. Evolutionary dynamics and significance of multiple subclonal mutations in cancer. DNA Repair (Amst). 08 2017;56:7–15. doi:10.1016/j.dnarep.2017.06.002

10. McCoy M, Yeang CH, Bahnassy S, Tam S, Riggins RB, Parashar D, and Beckman RA, Generalized Evolutionary Classifier for Evolutionary Guided Precision Medicine, JCO Precis Oncol 2025, 9:e2300714. Doi: 10.1200/PO.23.00714

11. Buyse M, Michiels S, Sargent DJ, Grothey A, Matheson A, de Gramont A. Integrating biomarkers in clinical trials. Expert Rev Mol Diagn. 2011; 11(2):171–82. doi:10.1586/erm.10.120

12. Beckman RA, Clark J, Chen C. Integrating predictive biomarkers and classifiers into oncology clinical development programmes. Nat Rev Drug Discov. 2011; 10(10):735–48. doi:10.1038/nrd3550

13. Freidlin B, McShane LM, Polley MY, Korn EL. Randomized phase II trial designs with biomarkers. J Clin Oncol. 2012; 30(26):3304–9. doi:10.1200/JCO.2012.43.3946.

14. Parashar D, Bowden J, Starr C, Wernisch L, Mander A. An optimal stratified Simon two-stage design. Pharm Stat. 2016;15(4):333–333. doi:10.1002/pst.1742.

15. Simon N, Simon R. Adaptive enrichment designs for clinical trials. Biostatistics. 2013; 14(4):613–25. doi:10.1093/biostatistics/kxt010

16. Zhang J, Cunningham J, Brown J, Gatenby R. Evolution-based mathematical models significantly prolong response to abiraterone in metastatic castrate-resistant prostate cancer and identify strategies to further improve outcomes. eLife 2022;11: e76284.

## Supplementary References

1. Baca OG, Scott TO, Akporiaye ET, DeBlassie R, Crissman HA (1985) Cell cycle distribution patterns and generation times of L929 fibroblast cells persistently infected with Coxiella burnetii. Infect Immun 47:366–369.

2. Baker FL, Sanger LJ, Rodgers RW, Jabboury K, Mangini OR (1995) Cell proliferation kinetics of normal and tumour tissue in vitro: Quiescent reproductive cells and the cycling reproductive fraction. Cell Prolif 28(1):1–15.

3. MITOPENCOURSEWARE (2005) Cell, tissue and tumor kinetics. Available at http://ocw.mit.edu

4. Haeno H, et al. (2012) Computational modeling of pancreatic cancer reveals kinetics of metastasis suggesting optimum treatment strategies. Cell 148:362–375.

5. Cervantes RB, Stringer JR, Shao C, Tischfield JA, Stambrook PJ (2002) Embryonic stem cells and somatic cells differ in mutation frequency and type. Proc Natl Acad Sci USA 99:3586–3590.

6. Zhang J, et al. (2012) A novel retinoblastoma therapy from genomic and epigenetic analyses. Nature 481:329–334.

7. Camps M, Naukkarinen J, Johnson BP, Loeb LA (2003) Targeted gene evolution in Escherichia coli using a highly error-prone DNA polymerase I. Proc Natl Acad Sci USA 100:9727–9732.b

8. Beckman, R. A., Schemmann, G. S., & Yeang, C. H. (2012). Impact of genetic dynamics and single-cell heterogeneity on development of nonstandard personalized medicine strategies for cancer. Proceedings of the National Academy of Sciences of the United States of America, 109(36), 14586–14591.

